# Body mass index and carpal tunnel syndrome: a Mendelian randomization study

**DOI:** 10.1101/19011536

**Authors:** Iyas Daghlas, Nathan Varady

**Affiliations:** Harvard Medical School, 25 Shattuck Street, Boston, MA, 02115, USA

**Keywords:** Mendelian randomization, carpal tunnel syndrome, body mass index, obesity, entrapment neuropathy

## Abstract

**Introduction:** We investigated whether higher body mass index (BMI) may causally increase risk of carpal tunnel syndrome using Mendelian randomization (MR).

**Methods:** The exposure included genetic instruments comprising independent variants associated with BMI (n=322,154). Associations of these variants with CTS were obtained from a genome-wide association study conducted in UK Biobank (UKB; 12,312 cases/389,344 controls) with replication in the FinnGen cohort (4,505 cases/86,854 controls). Causal effects were estimated using inverse-variance weighted regression, and sensitivity analyses probed robustness of results.

**Results:** A 1-standard deviation (4.7kg/m^2^) increase in genetically instrumented BMI increased the odds of CTS in UKB (OR 1.73, 95% CI 1.48-2.02, *P*=2.68×10^−12^; 16.8 additional cases per 1000 person-years [95% CI 11.0-23.5]). This effect was consistent in the replication sample and across sensitivity analyses.

**Discussion:** These data support a causal effect of higher BMI on susceptibility to CTS. Clinical investigations of weight loss for treatment of CTS may be warranted.

## Introduction

Carpal tunnel syndrome (CTS) is the most common entrapment neuropathy, with an estimated prevalence ranging from 1-5%^1^. CTS accounts for a large proportion of lost work time related to disabling injuries, and medical care for CTS has been estimated to cost more than $2 billion annually^2^. In light of this clinical and economic burden of disease, efforts have been made to delineate modifiable factors contributing to risk of CTS^3^.

Elevated body mass index (BMI), a measure of overall adiposity, has consistently emerged as a modifiable risk factor for CTS^3^. A meta-analysis of 58 observational studies related obesity to a doubling in odds of CTS^4^. However, given the observational nature of these studies, it is unclear whether this association is causal^5^. For instance, BMI is influenced by many underlying health conditions such as hypothyroidism^6^, which may confound the relationship of obesity with CTS. Establishing evidence for a causal effect of higher BMI on CTS would strengthen the case for investment in trials investigating weight loss for treatment of CTS.

Limitations to causal inference in observational research may be overcome by an instrumental variable method called ‘Mendelian randomization’ (MR)^7^. MR uses genetic variants as proxies for epidemiologic exposures (e.g. BMI) to test for causal effects on outcomes (e.g. CTS). This method leverages the random assortment of genetic variants at gametogenesis which, at the population level, randomly allocates individuals to different levels of any given heritable exposure. This random allocation substantially reduces confounding and minimizes the impact of reverse causality when testing epidemiologic relationships^5^. We thus leveraged MR to determine whether higher BMI causally increased CTS risk.

## Methods

### Overall study design

We implemented a two-sample MR study design. Two-sample MR uses genetic variants to estimate the causal effect of a given exposure on an outcome. Non-overlapping samples are used to measure the variant-exposure and variant-outcome effect. Three key assumptions must be met for unbiased causal effect estimation in MR: 1) genetic variants must be strongly associated with the exposure, 2) the association of the genetic variant with the exposure or outcome must not be confounded, and 3) the genetic variants must only affect the outcome through the exposure of interest^5^. These three assumptions are addressed in turn below.

### Exposures and mediators

To identify genetic instrumental variables proxying effects of BMI, we drew upon genome-wide association studies (GWAS) conducted by the Genetic Investigation of ANthropometric Traits (GIANT) consortium. This comprised a meta-analysis across 80 cohorts, collectively contributing data on up to 322,154 participants of European ancestry^8^. The per-allele effects of the variants on BMI are reported in relation to a 1-standard deviation (SD) increase in BMI, corresponding to 4.7 kg/m^2^. This approximately represents the transition from overweight to obesity.

As some observational evidence suggests that higher central adiposity may also independently increase CTS risk^4^, we tested waist-to-hip ratio adjusted for BMI (WHRadjBMI), a validated measure of central adiposity^9^, as an exposure in secondary analyses. The genetic association study for WHRadjBMI comprised a meta-analysis across 101 cohorts, collectively contributing data on up to 210,088 participants of European ancestry^9^. Here, a 1-SD change represents 0.08 units of WHRadjBMI. The genetic instruments for both BMI and WHRadjBMI have been validated in independent cohorts^10,11^.

We also tested whether the effects of BMI on CTS were attributable to the effects of BMI on blood lipids (low-density lipoprotein cholesterol (LDL-c), high-density lipoprotein cholesterol (HDL-c), and triglycerides) or on type 2 diabetes (T2D), both of which are influenced by BMI^12,13^ and are putative risk factors for CTS^14,15^. Although there are larger T2D GWAS available^31^, this dataset was selected to avoid sample overlap with UKB. Details on the genetic association studies for these mediators are provided in Table 1.

**Table 1.**
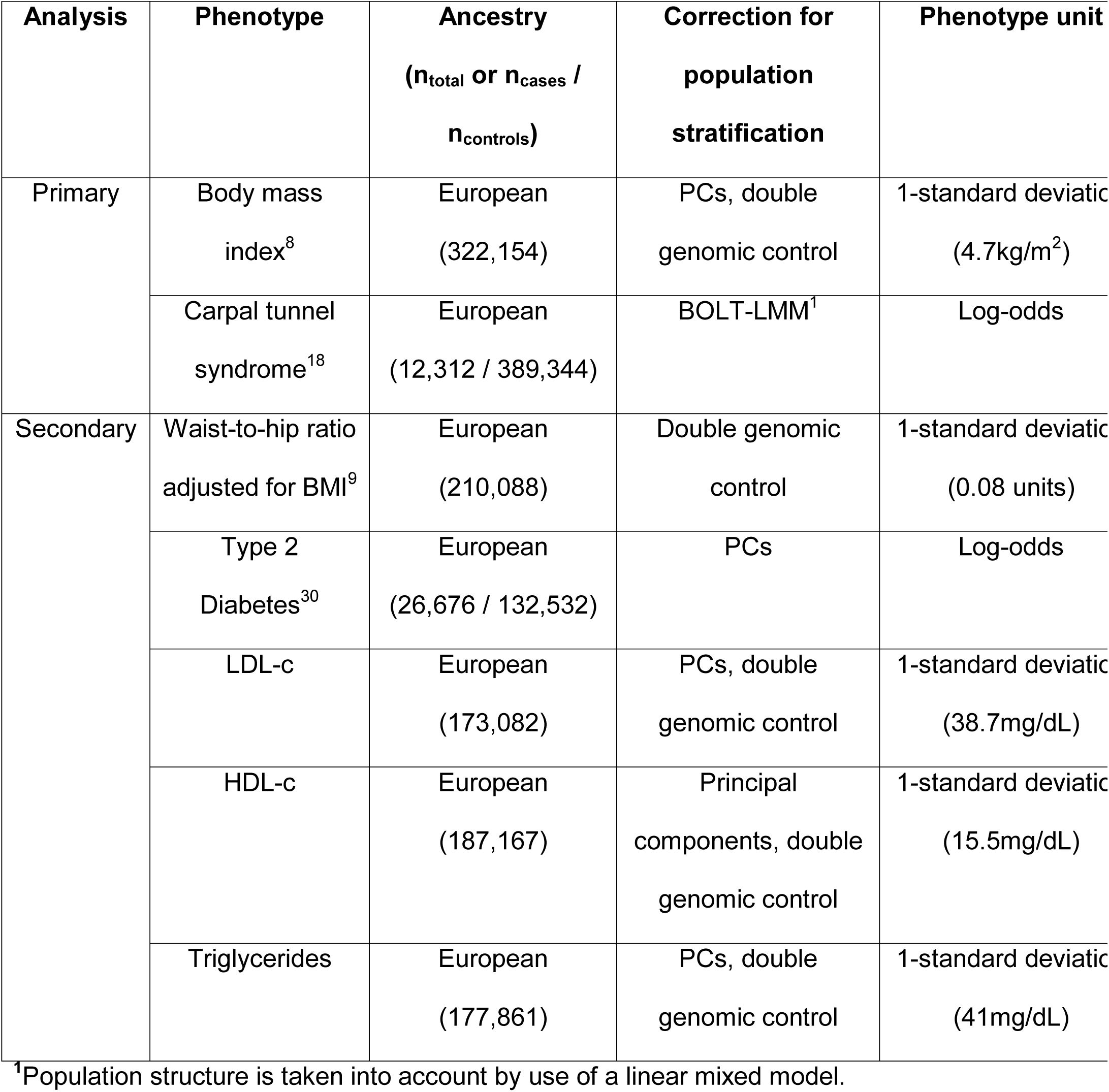
GWAS datasets contributing genetic association estimates for use in MR analysis. LDL-c: low-density lipoprotein cholesterol; HDL-c: high-density lipoprotein cholesterol; PCs: principal components of ancestry; T2D: type 2 diabetes.

### Outcome dataset

The outcome data source was a GWAS of carpal tunnel syndrome (CTS) conducted using the population-based UK Biobank (UKB) cohort. Details regarding the UKB cohort design and population have been previously reported^16^. Approximately 500,000 participants were recruited across 22 assessment centers from 2006-2010. At baseline assessment, the volunteers completed a standardized questionnaire and interview with a study nurse, and blood was collected for genotyping.

Cases of CTS were defined as UKB participants meeting any of the following criteria: self-reported history of CTS, ICD-10 code for CTS, operation code for carpal tunnel release or revision of carpal tunnel release, and self-reported history of carpal tunnel surgery. Identification of CTS cases in the UKB through routine health record data has been validated to have a high positive predictive value as compared to manual record review by experienced clinicians^17^. In total, 12,312 CTS cases and 389,344 controls of European ancestry with suitable genetic data were identified for analysis. The contribution of each data source to the total case count has been previously reported^18^, with the ICD-10 code G560 contributing the greatest number of cases (n=9,139). GWAS was conducted using BOLT-LMM to account for relatedness within the UKB cohort, with linear regression beta coefficients converted to log-odds using the prevalence of CTS in UKB^18^.

We sought to replicate the effect of BMI on CTS using publicly available CTS GWAS summary statistics from the FinnGen consortium. Details on genotyping and imputation are available online (see Data Links at end of manuscript). In brief, samples were genotyped with Illumina and Affymetrix arrays, imputed using the SISu population-specific reference panel, and analyzed using the SAIGE^19^ GWAS software to account for sample relatedness. CTS cases were exclusively identified using ICD codes (ICD-9: 3540; ICD-10: G560) contributed by nationwide Finnish registries. In total, there were 4,505 CTS cases and 86,854 controls.

### Selection of genetic instrumental variables and data harmonization

We extracted variants associated with BMI at genome-wide significance from the GWAS summary statistics file (p<5×10^-8^). This threshold corresponds to an F-statistic of 30, which is greater than the accepted cutoff of 10 to mitigate weak instrument bias, and hence satisfies the MR assumption that the genetic instrument has a strong association with the exposure (i.e. the ‘relevance assumption’)^5^. From this variant list, we identified variants also present in the CTS GWAS that could be matched on the basis of allele frequency, and harmonized the SNP-exposure associations with the SNP-outcome associations by matching effect alleles. To generate a set of independent genetic variants, we clumped this set of variants to a between-SNP linkage disequilibrium cutoff of r^2^ of 0.001 using the 1,000 Genomes European reference panel implemented in the TwoSampleMR package^20^. This approach for identifying variants for use as instrumental variables was taken for WHRadjBMI and each of the considered mediators.

This variant selection procedure identified 69 independent SNPs for use as instrumental variables for BMI. Using previously described methods for summarized data^21^, we determined that these variants collectively explained 2.27% of the variance in BMI. Using this variance estimate in the mRnd power calculator^22^, we determined that our study had 80% power to detect a causal OR of 1.17 for CTS. Notably, the percent variance explained informs the power, but not validity, of the MR analysis.

### Primary MR analyses

To estimate the causal effect of BMI on CTS risk, we regressed the SNP-CTS associations on the SNP-exposure associations, and weighted the effects by the inverse of the variance (IVW) of the SNP-CTS association under a random-effects model with the intercept constrained at the origin^23^. The estimated odds ratio approximates the relative risk given the population prevalence of CTS in UKB, which permitted conversion of effect estimates to absolute risk increases using the following equation^11^: absolute risk increase per 1-SD increase in exposure = [OR-1] x [absolute incidence]. Incidence estimates used for absolute risk calculations were obtained from a pooled analysis of US working populations, as part of the upper-extremity musculoskeletal disorder consortium (23 CTS cases per 1000 person-years^24^).

### Sensitivity analyses for horizontal pleiotropy

In addition to the assumption that the SNPs are strongly associated with the exposure of interest, there are two assumptions required for valid causal inference in Mendelian randomization. First, the genetic associations must not be confounded by factors that influence both genotype and the phenotype of interest. This is unlikely to be a major bias given the random allocation of genetic variants at gametogenesis^5,25^ and our use of data from GWAS that have carefully controlled for population stratification (Table 1). Population stratification is defined as systematic differences in allele frequencies between cases and controls which are attributable due to differences in ancestry rather than direct effects of genes on disease. Second, the genetic variants associated with the exposure must not influence the outcome through pathways independent of the exposure of interest. This is the strongest assumption of MR, and is violated in the setting of horizontal pleiotropy. We therefore conducted a range of sensitivity analyses that produce unbiased estimates under various forms of pleiotropy. The robust, heterogeneity-penalized IVW method down weights the influence of heterogeneous, potentially pleiotropic, SNPs MR-Egger models an intercept term in the IVW regression that captures potential unbalanced horizontal pleiotropy^27^. In addition to producing an effect estimate, the MR Egger intercept estimate provides a measure of unbalanced horizontal pleiotropy. The weighted median estimator provides consistent causal effect estimation when up to 50% of the variants are pleiotropic^28^. Finally, we utilized the Mendelian Randomization Pleiotropy RESidual Sum and Outlier (MR-PRESSO) method to detect and remove potentially pleiotropic variants^29^. We considered consistent effects across sensitivity analyses to strengthen evidence for causality.

### Assessment of mediation through cardiometabolic risk factors

We aimed to determine whether any of the observed effect of BMI on CTS may be attributable to its effects on T2D or blood lipids. To do so, we first tested for effects of the putative mediators with CTS in univariable MR. Consistent with prior work utilizing binary exposures in MR, the effect of diabetes liability on CTS was scaled to represent a doubling in the odds of diabetes^32^. For mediators with an identified effect in univariable MR, we conducted regression-based multivariable^33^ MR accounting for the effect of BMI on CTS through the mediator (e.g. BMI -> T2D -> CTS). We considered an attenuation in the BMI coefficient to be consistent with mediation through the risk factor of interest. As previously recommended for mediation analysis involving dichotomous outcomes in MR ^34^, we report the total (from univariable MR) and direct (from multivariable MR) effects of BMI.

### Software and statistical analysis

All analyses were conducted using R version 3.5.0. We utilized the TwoSampleMR^20^ and MendelianRandomization packages. All statistical tests were two-sided.

### Standard Protocol Approvals, Registrations, Patient Consents, and Data availability

This study used precompiled, publically available, de-identified data provided by genetic consortia and therefore did not require IRB approval. Informed consent was obtained from all participants in contributing studies. All data used in this study are publicly available at the links provided at the end of the manuscript.

## Results

### Mendelian randomization supports a causal effect of BMI on risk of CTS

A 1-SD increase in BMI increased risk of CTS (OR 1.73, 95% CI 1.48-2.02, *P*=2.68×10^−12^; Figure 1). This effect corresponds to an absolute risk increase of 16.8 additional CTS cases per 1000 person years [95% CI 11.0-23.5]. This effect replicated in the independent FinnGen cohort (OR 1.62, 95% CI 1.30-2.02, *P*=1.58×10^−5^). In contrast, a 1-SD increase in WHRadjBMI was not associated with CTS in UKB (OR 1.01, 0.76-1.34, *P*=0.97; Table 1).

**Figure 1.**
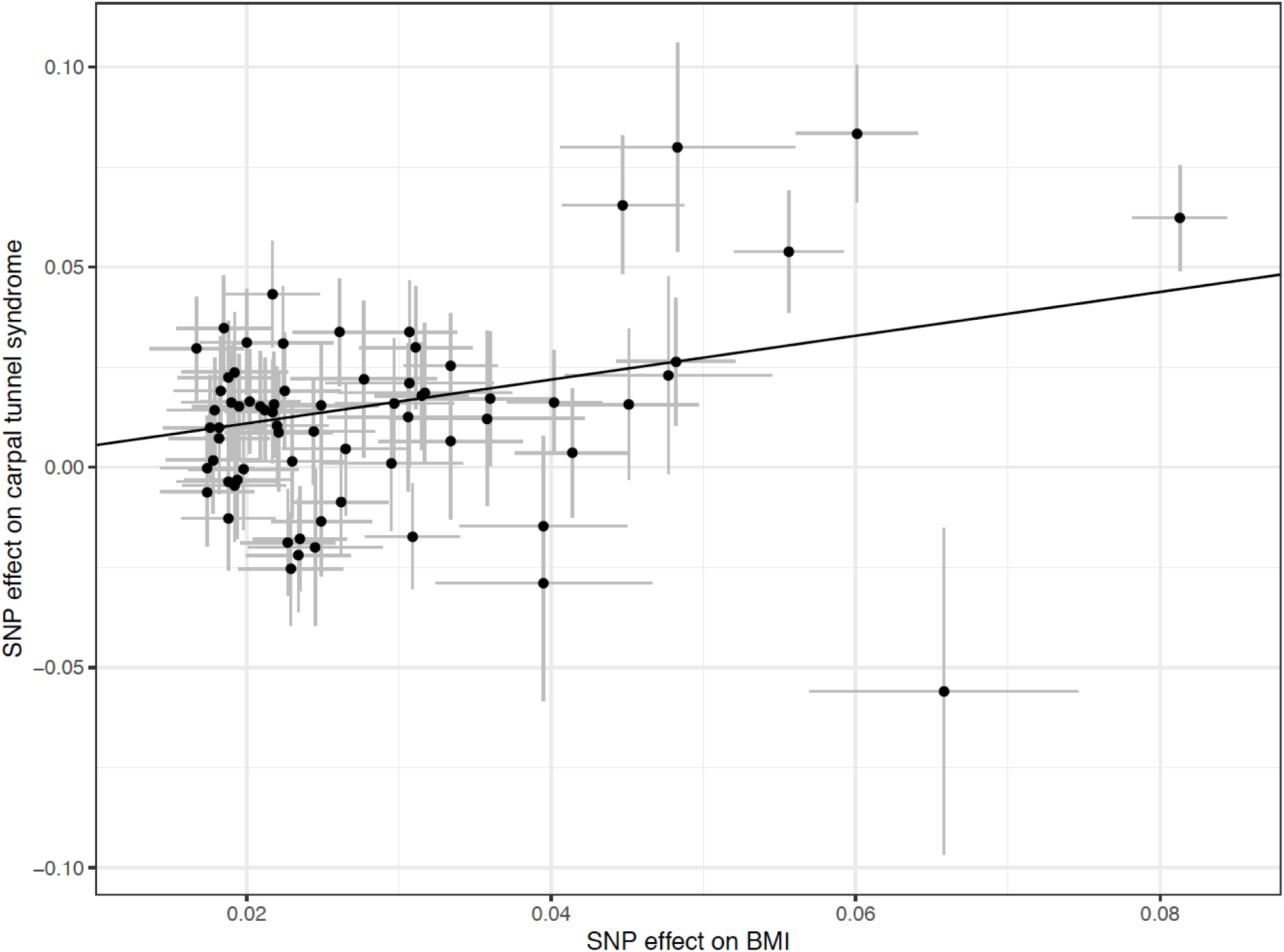
Scatter-plot of BMI-increasing variants and their effects on carpal tunnel syndrome (CTS). The x-axis shows the variant effects on BMI (standard deviation units), and the y-axis shows the variant effects on CTS (log-odds). The regression line represents the inverse-variance weighted regression of CTS on BMI, with the slope representing the estimate of the casual effect.

### Sensitivity analyses do not indicate that the causal effect of BMI on CTS is driven by horizontal pleiotropy

As there was evidence of heterogeneity in the causal effect of BMI on CTS (*P*=6.7×10^−4^), we conducted sensitivity analyses to assess whether the effect of genetically predicted BMI on CTS may be biased by pleiotropy. First, the MR Egger intercept, a measure of unbalanced horizontal pleiotropy, did not differ from the null (Beta = −0.01, *P*=0.21). Second, all three models used in sensitivity analyses provided consistent results (Figure 2). Third, the MR-PRESSO method did not detect any potentially pleiotropic outlier variants in the BMI instrument. Thus, the primary MR result is unlikely to be biased by horizontal pleiotropy.

**Figure 2.**
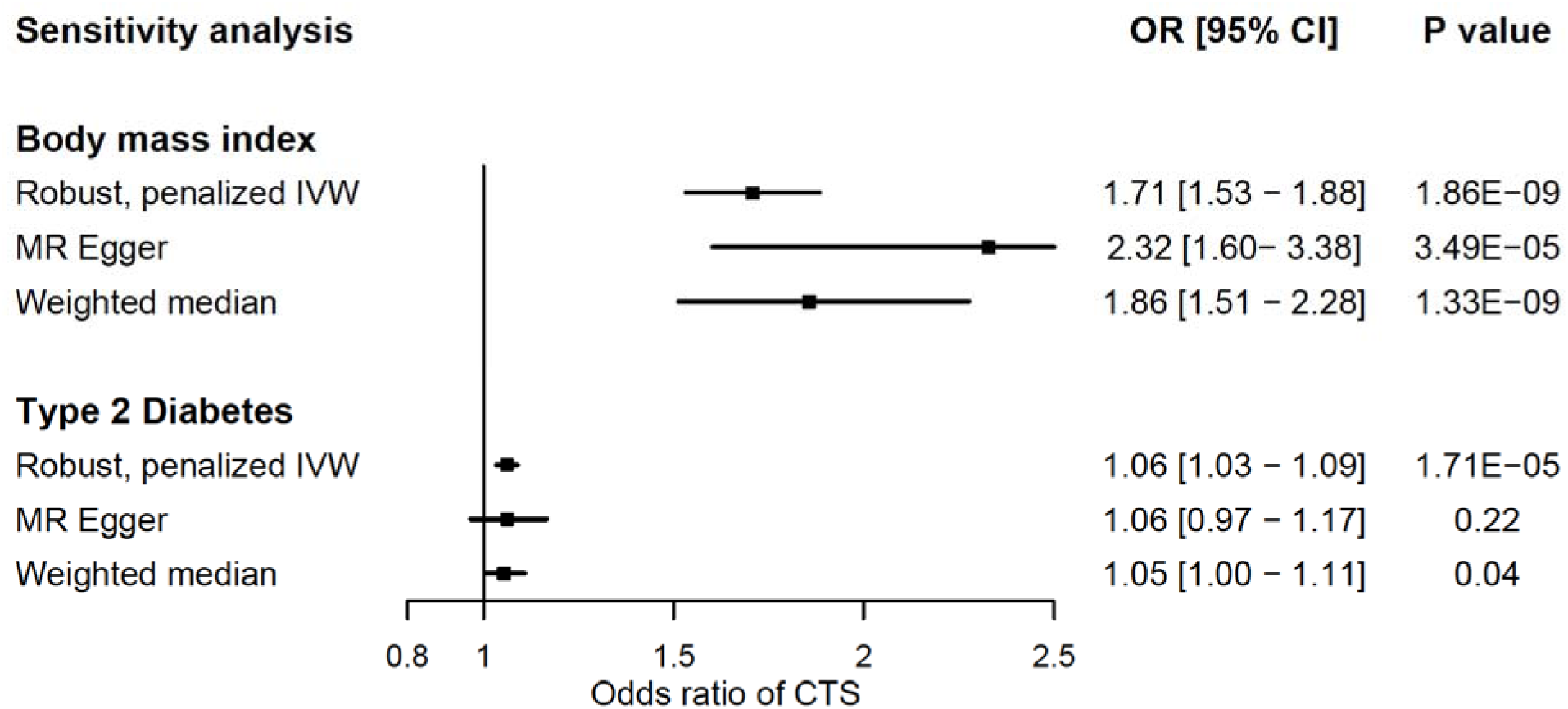
Mendelian randomization sensitivity analyses for the effect of genetically instrumented BMI (per standard deviation increase) and T2D^a^ liability on odds of carpal tunnel syndrome (CTS). Bars represent 95% confidence intervals. BMI: body mass index; CI: confidence interval; OR: odds ratio; SD: standard deviation; T2D: type 2 diabetes ^a^Units of the exposure are a doubling in the odds of T2D.

### Type 2 diabetes partially mediates the effect of BMI on CTS

Having established a robust effect of higher BMI on increased risk of CTS, we tested whether any portion of this total effect was attributable to knock-on effects of higher BMI on blood lipids or diabetes risk. Liability to T2D increased risk of CTS (OR 1.08, 95% CI 1.03-1.11, P=5.20×10^−5^), with consistent effects in pleiotropy-robust sensitivity analyses (Figure 2). The effect of higher BMI on CTS was partially attenuated in multivariable MR when accounting for mediation through T2D (1.53, 95% CI 1.38−1.68, *P*=2.85×10^−8^; Table 1). In contrast, the effects of blood lipids on CTS were null, suggesting that any effect of BMI on lipids is not responsible for its role in causing CTS.

**Table 2.**
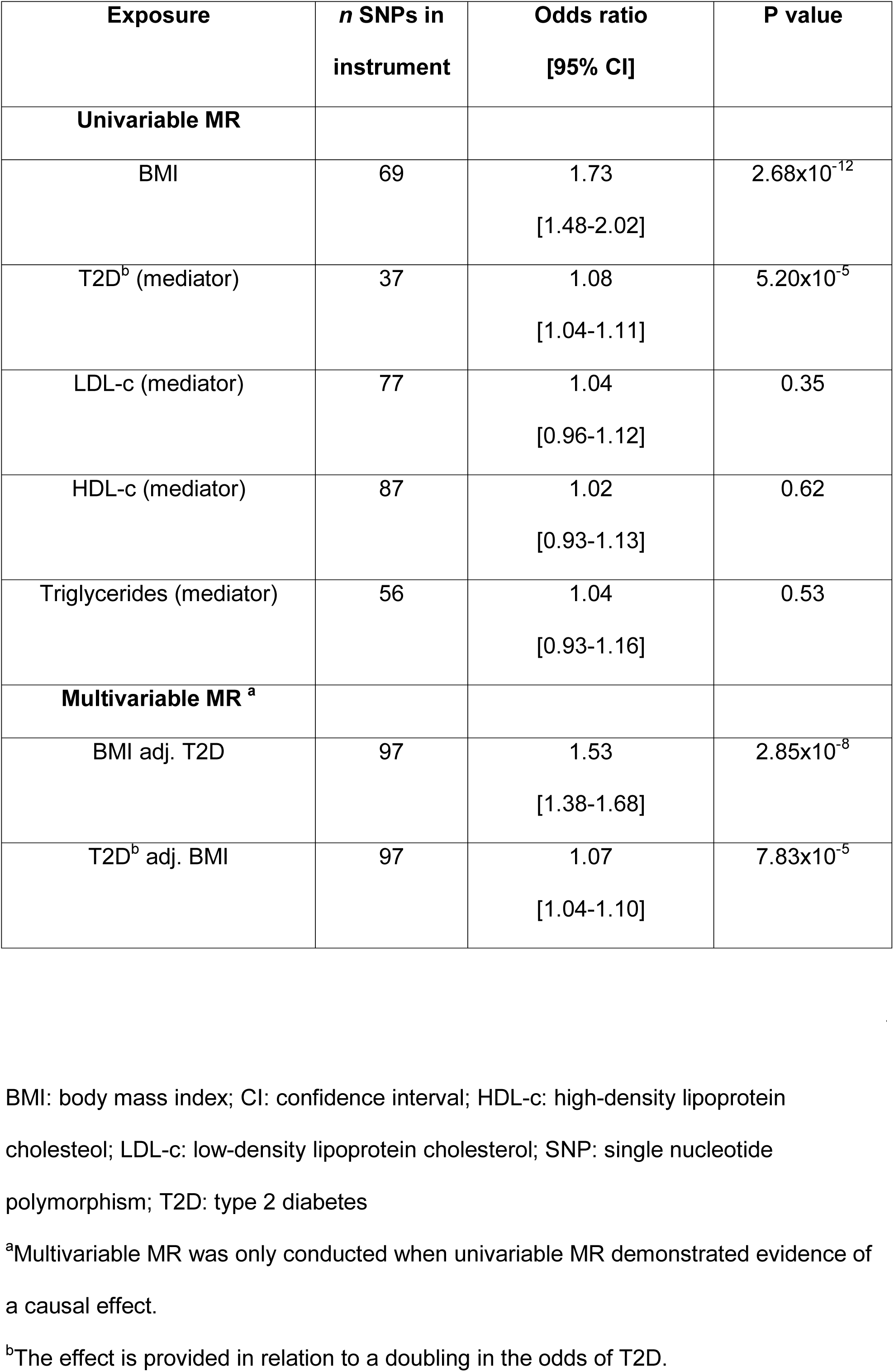
Inverse-variance weighted and multivariable MR effects of BMI and mediators on carpal tunnel syndrome in UK Biobank. LDL-c: low-density lipoprotein cholesterol; HDL-c: high-density lipoprotein cholesterol; PCs: principal components of ancestry; T2D: type 2 diabetes.

## Discussion

While numerous observational studies have found elevated BMI to be associated with CTS^4^, whether BMI plays any causal role in CTS risk has remained unknown. Using a pseudo-randomized study design, we found strong evidence for a causal link between higher BMI and increased risk of CTS. Furthermore, we replicated these results in an independent sample and found the effects to be consistent across a range of sensitivity analyses assessing potential violations of the MR assumptions. We also provide mechanistic evidence for how BMI may exert this effect on CTS, namely through its role in causing T2D. Taken together, these results substantially enhance our understanding of BMI as a key CTS risk factor and have a number of implications in the prevention and treatment of CTS.

The most important finding of this study was the strong evidence that higher BMI causally increased risk of CTS. The present analysis is able to overcome potential biases of prior observational studies by utilizing genetic variants as proxies for higher BMI. Specially, given the random allocation of genetic variants at gametogenesis^5,36^ and the study-specific control for confounding by population stratification, our results are unlikely to be biased by confounding or reverse causality. Consequently, these results greatly strengthen the case for investment in trials investigating weight loss for treatment of CTS. Furthermore, in considering the increasing rates of obesity in adults^37^, our results may partly explain the rising CTS incidence^38^. Population-level efforts to address the obesity epidemic may therefore have substantial benefits in stemming the increase in rates of CTS.

Our results inform the mechanistic understanding of how BMI influences CTS by identifying mediation through T2D. Type 2 diabetes is observationally associated with CTS^14^, potentially through increased oxidative stress and extracellular protein glycation^4^. In univariable MR we found evidence to suggest that this effect is causal, with no evidence for bias by horizontal pleiotropy. In multivariable MR we found evidence suggesting that part of the effect of BMI on CTS is accounted for by T2D. However, the direct effect of BMI on CTS independently of T2D remained significant (OR 1.53). Therefore, obese populations remain at elevated risk for CTS regardless of their T2D status. In contrast, other dimensions of metabolic health including central adiposity proxied by WHRadjBMI and circulating lipids did not have a causal relationship with CTS. Thus, apart from T2D, these results do not support cardiometabolic risk as a strong contributor to CTS. Furthermore, these results do not implicate LDL-c lowering as a causal risk factor for peripheral neuropathies, as has previously been suggested^39^. Observational associations relating lipids to risk of CTS may have been driven by residual confounding, potentially through BMI or T2D^15^.

Having established a likely role for higher BMI in the etiology of CTS, it is plausible that weight loss may be a feasible strategy for prevention or treatment of CTS. Indeed, a longitudinal study of patients undergoing bariatric surgery found lower rates of CTS after bariatric surgery^40^. However, the mechanisms by which this effect of BMI may operate independently of T2D remains speculative. A direct compressive effect of adipose tissue within the carpal tunnel, or of increased hydrostatic pressure throughout the carpal canal due to surrounding adipose tissue^41^, has been suggested^4^. Alternatively, the link may be explained through increased endoneurial edema^42^ or through increased rates of hand osteoarthritis^43^ in obese populations^44^.

The major strength of this analysis is the use of MR, with consistent results across sensitivity analyses. The use of non-overlapping samples for measurement of genetic associations with the exposure and outcome avoids biases related to sample overlap^45^. Finally, independent replication lends further support to the robustness of the reported association. This is particularly relevant to UKB, which may be biased due to the relative healthiness of the study participants^46^. There are limitations to acknowledge. First, these findings relate to CTS etiology rather than progression or severity. Testing such effects would necessitate a GWAS of clinical progression, severity, or of measures derivedDaghlas 15 BMI and carpal tunnel syndrome from electrodiagnostic testing^47^. Second, horizontal pleiotropy may bias the causal effects^5^; however, our results were consistent across a range of sensitivity analyses suggesting this is an unlikely explanation. Third, CTS may be under-reported in UKB, which may result in an underestimation of the true causal effect in our analyses (i.e. bias towards the null). Fourth, we did not conduct analyses stratified by sex due to lack of available sex-stratified GWAS. Indeed, it is plausible that the effect of higher body mass index differs by sex, as female sex is a risk factor for CTS^44^.

In conclusion, these results are consistent with a causal effect of higher BMI on increased risk of CTS. Clinical investigations of weight loss in the management of CTS may be warranted.

## Data Availability

All summary statistics used for the present analysis are publically available.

## Data links

CTS GWAS: https://static-content.springer.com/esm/art%3A10.1038%2Fs41467-019-08993-6/MediaObjects/41467_2019_8993_MOESM6_ESM.txt

BMI and WHRadjBMI GWAS: https://portals.broadinstitute.org/collaboration/giant/index.php/GIANT_consortium_data_files

GLGC GWAS: http://lipidgenetics.org/

DIAGRAM GWAS: https://www.diagram-consortium.org/

FinnGen GWAS: https://finngen.gitbook.io/finngen-documentation/-LvQ4yR2YFUM5eFTjieO/

mRnd power calculator: http://cnsgenomics.com/shiny/mRnd/)

## Acknowledgements

We are thankful to the authors of the CTS GWAS for making their data available, as well as the following GWAS consortia for providing publically available data: GIANT, GLGC, and DIAGRAM. We want to acknowledge the participants and investigators of the UK Biobank and FinnGen studies.

